# Scoping review of Japanese encephalitis virus transmission models

**DOI:** 10.1101/2024.07.08.24310107

**Authors:** Troy A. Laidlow, Erin S. Johnston, Ruth N. Zadoks, Michael Walsh, Mafalda Viana, Kerrie E. Wiley, Balbir B. Singh, Francesco Baldini, Himani Dhanze, Cameron Webb, Victoria J. Brookes

## Abstract

Japanese encephalitis virus (JEV) causes approximately 100,000 clinical cases and 25,000 deaths annually worldwide, mainly in South-East Asia and the Western Pacific and mostly in children. JEV is transmitted from competent hosts to humans through the bite of mosquitoes, and the abiotic environment, such as seasonal rainfall, influences transmission. Transmission models have an important role in understanding disease dynamics and developing prevention and control strategies to limit the impact of infectious diseases. Our goal was to investigate how transmission models capture JEV infection dynamics and their role in predicting and controlling infection. This was achieved by identifying published JEV transmission models, describing their features, and identifying their limitations, to guide future modelling. A PRISMA-ScR guided scoping review of peer-reviewed JEV transmission models was conducted. Databases searched included PubMed, ProQuest, Scopus, Web of Science and Google Scholar. Of 881 full text papers available in English, 29 were eligible for data extraction. Publication year ranged from 1975 to 2023. The median number of host populations represented in each model was 3 (range: 1–8; usually humans, mosquitoes and pigs). Most (72% [n=21]) models were deterministic, using ordinary differential equations to describe transmission. Ten models were applied (representing a real JEV transmission setting) and validated with field data, while the remaining 19 models were theoretical. In the applied models, data from only a small proportion of countries in South-East Asia and the Western Pacific were used. Limitations included gaps in knowledge of local JEV epidemiology, vector attributes and the impact of prevention and control strategies, along with a lack of model validation with field data. The lack and limitations of models highlight that further research to understand JEV epidemiology is needed and that there is opportunity to develop and implement applied models to improve control strategies for at-risk populations of animals and humans.

## 1. Introduction

Japanese encephalitis (JE) is the leading form of human acute viral encephalitis in Asia and the Pacific, and it is estimated that more than 1.5 billion people live in areas suitable for endemic JE (Erlanger et al. 2009; Moore 2021). JE is caused by Japanese encephalitis virus (JEV), a zoonotic mosquito-borne orthoflavivirus. In reported human cases, estimated case fatality approaches 30% and of those who survive, an estimated 46% suffer permanent neurological sequelae (Cheng et al. 2022). Studies of the global burden of JE estimated 100,308 cases (95% CI: 61,720–157,522) and 25,125 deaths (95% CI: 14,550–46,031) in 2015 with most cases occurring in children aged 0–14 years (incidence: 5.4 per 100,000) (Quan et al. 2020; Campbell et al. 2011). However, it is likely that reported case numbers are inaccurate due to inadequate data collection and diagnosis, or attributed incorrectly due to cross-reactivity of serological tests with other flaviviruses (Maeki et al. 2019).

It is generally accepted that JEV is transmitted from a wild reservoir host (such as ardeid birds, i.e., herons and bitterns) or an amplifying host (such as wild or domestic pigs) to humans through the bite of mainly *Culex* spp. mosquitoes (Van Den Hurk, Ritchie, and Mackenzie 2009; De Wispelaere, Desprès, and Choumet 2017; Faizah et al. 2020). An experimental study showed that vector-free transmission between pigs can occur via oronasal infection, however this is yet to be reported under field conditions (Ricklin et al. 2016). The epidemiology of JEV might differ between regions based on variation in the infection ecology, particularly the diversity, abundance and composition of animal (non-human mammals and birds) and vector communities, and the circulating JEV genotype. Chicks and ducklings develop viremia (Page et al. 2014) and JEV outbreaks have been strongly associated with chicken density (Walsh et al. 2022), but further investigations are required to determine if poultry are competent hosts. Cattle, horses and dogs, like humans, have insufficient viremia to infect susceptible mosquitoes and are noncompetent hosts for JEV (Boyer et al. 2021; Yang et al. 2008). Mosquito host feeding preferences and changes in the ratio of competent hosts to noncompetent hosts – for example, in communities with livestock – can impact JEV transmission (Marini et al. 2017). Two epidemiological patterns of JEV have been described: endemic activity in tropical regions and epidemic activity in temperate and subtropical regions (Van Den Hurk, Ritchie, and Mackenzie 2009).

Disease transmission models are data driven mathematical approaches to understand the parameters responsible for the dynamics of pathogen transmission and to assess the strategic responses to disease risk (Becker et al. 2021). They can incorporate environmental, host and vector data to determine factors that influence the size and duration of outbreaks for vector-borne diseases such as dengue (Ogunlade et al. 2023) and malaria (Mandal, Sarkar, and Sinha 2011). These models have been valuable in the planning and evaluation of interventions, determining optimal prevention and control strategies, and predicting the expected course of disease events (Garnett et al. 2011). However, both model development and assessment can vary widely and, therefore, so can model accuracy and reliability. This can be due to epistemic uncertainty (imprecise knowledge of parameters), aleatoric uncertainty (due to randomness) (Penn et al. 2023), or to existing beliefs that influence model assumptions and interpretation of results (Garnett et al. 2011), all of which can hinder the appropriate generation and use of model outputs, especially for decision making.

In this scoping review, we aimed to examine how disease transmission models capture the dynamics of JEV infection and their use in prediction, prevention, and control of JEV spread. To achieve this, we collated and described peer-reviewed information in which models of JEV transmission in populations were developed or implemented. Models of JEV transmission were defined as those that made explicit hypotheses about the biological mechanisms that drive JEV infection dynamics in host and/or vector populations. We aimed to provide a baseline of current knowledge and knowledge gaps regarding JEV model development and parameterisation, host and vector population structures, and virus transmission between hosts. The findings of this review provide a foundation for the development of improved models of JEV transmission to support JE prevention and control.

## 2. Method

### 2.1 Protocol

This scoping review was conducted according to the Preferred Reporting Items for Systematic reviews and Meta-Analyses extension for Scoping Reviews (PRISMA-ScR) guidelines (Tricco et al. 2018). The objective was to collate and describe peer-reviewed information in which models of JEV transmission in populations were developed or used.

The review protocol comprised three levels: Level 1; screening on title and abstract; Level 2, screening on full record; Level 3 data extraction. The web-based review platform Sysrev (Bozada et al. 2021) was used for Levels 1 and 2 and a spreadsheet in Google Sheets (Google 2024) was used for Level 3.

We use the term ‘record’ to describe any bibliography citation captured in the searches. We use the term ‘model’ to describe a disease transmission model that was either developed or implemented to describe or quantify the transmission of JEV in populations.

A total of 11 reviewers participated in the scoping review. Reviewers were selected based on their knowledge of JEV or disease transmission modelling and/or their experience in performing scoping reviews.

### 2.2 Eligibility

Records were eligible for inclusion if they were peer-reviewed literature, including peer-reviewed conference proceedings, for which the full text was available, published in English, in any year, and from any country, and contained primary research of interest in which a model of JEV transmission was developed or implemented. Models could range from representation of transmission in one host population to models that explicitly represented the spatio-temporal variability and heterogeneous contact structures in multiple populations.

Theses, dissertations, and pre-prints were excluded. Records which described JEV statistical models (for example, inferential models that aimed to identify and predict spatio-temporal occurrence based on risk factors or time-series models) were excluded.

### 2.3 Information sources and search strategy

The literature search was conducted in January 2023, using the following combination of search terms:

i. “Japanese encephalitis” OR JEV,
ii. AND: model,
iii. AND: spread OR transmission,
iv. time frame: all,
v. language: “English”.

Four electronic databases were searched: PubMed, ProQuest, Scopus, and Web of Science (All databases) to provide a comprehensive search across various disciplines. A literature search, using the same criteria, was conducted via the Google Scholar search engine, in which the first 100 results were screened. All records were exported into the citation manager software Endnote, and duplicates were removed. Records were then uploaded to Sysrev for Level 1 screening.

### 2.4 Selection of relevant records – Level 1 and 2

During Level 1 (screening on title and abstract), two reviewers assessed each record. To maximise the sensitivity of identification of relevant records, records progressed to Level 2 if either reviewer assessed that the record might be eligible.

An agreement test was conducted prior to screening at Level 2 (screening on the full record), in which five reviewers screened the same randomly selected 20 records. Conflicting opinion about inclusion or exclusion of records were discussed to achieve agreement between reviewers and to refine and improve the clarity of the questions at each level (Table S1).

During Level 2, two reviewers initially assessed each record. Records were only included for charting in Level 3 if there was agreement that the record met the eligibility criteria between at least 2 reviewers. Conflicts of opinion were resolved via discussion and if required, consultation with a third reviewer.

### 2.5 Data items and charting process – Level 3

An agreement test was conducted prior to screening at Level 3, in which reviewers screened the same randomly selected five records. Conflicting opinion about inclusion or exclusion of records were discussed to achieve consensus between reviewers and to refine and improve the clarity of data extraction at Level 3 (Table S1).

During Level 3, two reviewers initially extracted data from each record. Conflicting opinions about extracted data were discussed between each record’s pair of reviewers and, if needed, a third reviewer to determine an agreement prior to synthesis of the extracted data.

Data items that were extracted included the year of publication, the type of modelling method and objective of the model (for example, to estimate the likely impact of available interventions), and if the model was applied (reflected a real JEV transmission setting), or theoretical. If the model had been applied using field data, the location of the data origin was also included. Regions were defined using the World Health Organization regions. All information incorporated into each model was also recorded and comprised vector and host species, weather variables, control and prevention strategies, validation strategies, and sensitivity analyses.

Data that were extracted about vector and host species included the number and species of populations used in the model, the compartments used to describe the structure of each population (for example, susceptible-infected-recovered [SIR]), and the parameters used to describe change between compartments of a population and infection transmission within the model.

Weather data extracted included spatial, seasonal and temporal variations of rainfall, temperature, and humidity, and the impact of weather variability on JEV transmission.

Control and prevention strategy data included the type of strategy used and its impact on JEV transmission, and how the strategy was incorporated in the model (for example, a parameter used to decrease total vector abundance in the event of vector control).

### 2.6 Identification of additional and missing records

The titles of references in the bibliography of two records that were retained for data analysis in Level 3 were checked to identify any records missed by the search strategy. If records were potentially relevant to the study, they were included in the review process using the same methods as records identified in the initial search.

A weekly literature search alert was created using the same four electronic databases with the same combination of search terms to monitor new studies being published after the initial literature search was conducted. If newly published records identified by the search alert were potentially relevant to the study, they were included in the review process using the same methods as records identified in the initial search. The search alert ceased in April 2024.

## 3. Results

### 3.1 Screening

Our search identified 881 records. Following removal of 309 duplicates, 572 records remained for screening (Figure 1). During Level 1 screening, 458 records were excluded, leaving 114 records for screening of full text (Level 2). Records were most commonly excluded because they were not relevant to JEV transmission modelling or were not primary literature. Overall, 29 records were included for data charting and synthesis in Level 3 (Table S1).

**Figure 1:**
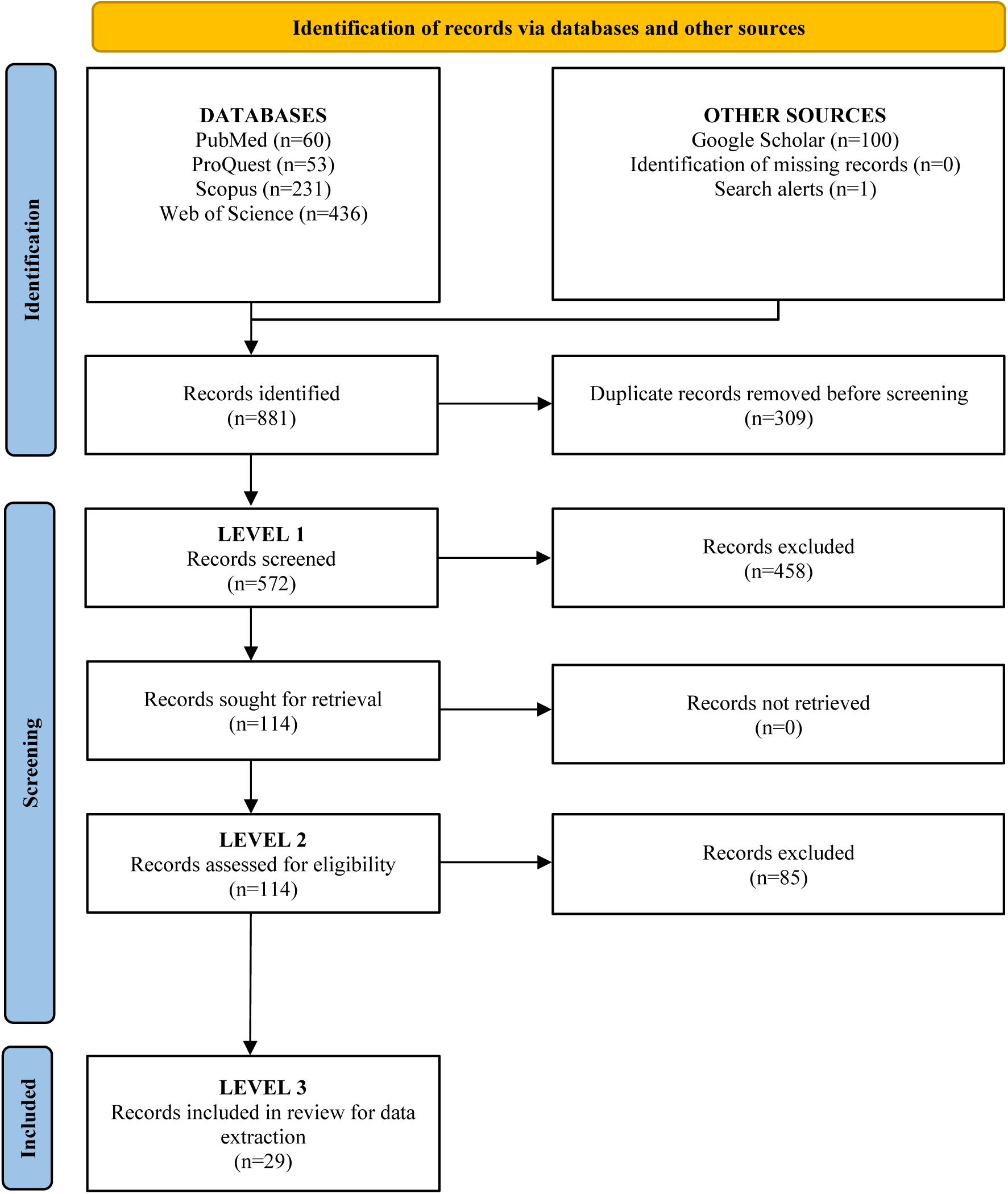
Diagram of the flow of records through the levels of a scoping review of Japanese encephalitis virus transmission models.

### 3.2 Data charting

All 29 records included in Level 3 described single models. Six records were published in peer-reviewed conference proceedings and the remainder were published in 22 peer-reviewed journals (Figure S1). Records were published from 1975 to 2023 (inclusive) (Figure S2).

Of the 29 models, 34% (n=10) were applied to real JEV transmission settings using data such as JE incidence and pig abundance and distribution. The most frequently represented region for these models was the Western Pacific (n=6) with models based in Cambodia, China, Philippines and Taiwan. Other locations included Bangladesh, French Overseas Département of Réunion, India, Japan, Thailand and United States of America (Figure S3).

### 3.3 Aims of models

Fifteen of the models aimed to draw general conclusions about JEV transmission dynamics, such as determining equilibrium points or reproductive numbers, and in some cases, conducting stability or sensitivity analyses related to these measures (Baniya and Keval 2021a, 2021c, 2020b; De et al. 2016; Diallo et al. 2018; Dwivedi, Keval, and Baniya 2022; Ghassabzade and Bagherpoorfard 2021; Ghosh and Tapaswi 1999; Goswami 2022; Kalita and Devi 2020a, 2020b; Mukhopadhyay and Tapaswi 1994; Panja, Mondal, and Chattopadhyay 2016; Tapaswi, Ghosh, and Mukhopadhyay 1995; Wada 1975). Eight models investigated the impact of various factors influencing host and vector species on the risk of JE in humans (Baniya and Keval 2020a, 2021b; Ladreyt, Chevalier, and Durand 2022; Ladreyt et al. 2023; Naresh and Pandey 2009; Ndaïrou, Area, and Torres 2020; Sota and Mogi 1989; Zahid and Kribs 2021). Four models aimed to describe, understand and predict JE incidence (Riad et al. 2017b, 2017a, 2019; Zhao et al. 2018). Lastly, two models aimed to describe the effects of interventions on human, animal-reservoir, or vector populations (Khan et al. 2014; Kharismawati and Fatmawati 2019).

### 3.4 Model structures

Twenty-two models were deterministic. Of these, 21 were implemented using continuous time, ordinary differential equations (Baniya and Keval 2021a, 2021c, 2020b, 2020a, 2021b; De et al. 2016; Diallo et al. 2018; Dwivedi, Keval, and Baniya 2022; Ghosh and Tapaswi 1999; Goswami 2022; Khan et al. 2014; Kharismawati and Fatmawati 2019; Ladreyt, Chevalier, and Durand 2022; Ladreyt et al. 2023; Mukhopadhyay and Tapaswi 1994; Naresh and Pandey 2009; Ndaïrou, Area, and Torres 2020; Panja, Mondal, and Chattopadhyay 2016; Sota and Mogi 1989; Tapaswi, Ghosh, and Mukhopadhyay 1995; Zahid and Kribs 2021) and one was implemented using discrete time, difference equations (Wada 1975). Four models were stochastic. Of these, two were implemented using continuous time, ordinary differential equations (Riad et al. 2017a; Zhao et al. 2018) and two were implemented using discrete time, difference equations (Riad et al. 2017b, 2019). The remaining three models were statistically converted models: two models were converted from deterministic to stochastic using geometric Brownian motion (Kalita and Devi 2020a, 2020b) and one converted a deterministic model implemented with ordinary differential equations to a deterministic fractional-order model (Ghassabzade and Bagherpoorfard 2021).

Two models simulated co-infection of the human population (JEV with either *Leptospira* spp. or dengue virus) (Dwivedi, Keval, and Baniya 2022; Zahid and Kribs 2021).

One model followed a single population of feral pigs across three spatial locations, representing each individual animal within a connected network (Riad et al. 2017a).

Models most commonly represented three populations — humans, vectors and an animal-reservoir (i.e., non-human mammal or bird) — but the number of populations in a model ranged from 1 to 8. A human population was represented in 24 models, a vector population in 21 models and at least one animal-reservoir population in 26 models.

#### 3.4.1 Human population

Compartments used in models to reflect different human infection categories and the transition of the human population over time were Maternal (M), Vaccinated (V), Susceptible (S), Exposed (E), Infected (I), and Recovered (R) (Table S2 for description of compartments). Models described the natural history of JEV in humans using nine different model structures and the most common was SIRS (n=7) (Table1).

**Table 1:** Number and types of model structures for human populations [*Maternal (M), Vaccinated (V), Susceptible (S), Exposed (E), Infected (I), and Recovered (R)*].

| Model structure |  | Citation |
| --- | --- | --- |
| SIRS | (n=7) | (Baniya and Keval 2021a; Ghosh and Tapaswi 1999; Goswami 2022; Kharismawati and Fatmawati 2019; Mukhopadhyay and Tapaswi 1994; Panja, Mondal, and Chattopadhyay 2016; Tapaswi, Ghosh, and Mukhopadhyay 1995) |
| SIS | (n=4) | (Ghassabzade and Bagherpoorfard 2021; Kalita and Devi 2020a; Naresh and Pandey 2009; Ndairou, Area, and Torres 2020) |
| VSIS | (n=4) | (Baniya and Keval 2021c, 2020a, 2021b; Kalita and Devi 2020b) |
| SEIR | (n=3) | (Ladreyt et al. 2023; Riad et al. 2017b, 2019) |
| VSIRS | (n=2) | (Baniya and Keval 2020b; De et al. 2016) |
| I | (n=1) | (Zhao et al. 2018) |
| MSEIR | (n=1) | (Ladreyt, Chevalier, and Durand 2022) |
| SIR | (n=1) | (Zahid and Kribs 2021) |
| VSIR | (n=1) | (Dwivedi, Keval, and Baniya 2022) |

#### 3.4.2 Vector population

Compartments used in models to reflect different vector infection categories and the transition of the vector population over time were Aquatic (A), Susceptible (S), Exposed (E), and Infected (I) (Table S2 for description of compartments). Mosquitoes were described as the main vector; however, details such as mosquito species and preferred habitat (e.g., water source and vegetation type) were not included in any models. Models described the natural history of JEV in vectors using four different model structures and the most common was SI (n=14) (Table 2).

**Table 2:** Number and types of model structures for vector populations [*Aquatic (A), Susceptible (S) Exposed (E), and Infected (I)*].

| Model structure |  | Citation |
| --- | --- | --- |
| SI | (n=14) | (Baniya and Keval 2021a, 2021c, 2020b, 2020a, 2021b; De et al. 2016; Dwivedi, Keval, and Baniya 2022; Goswami 2022; Kharismawati and Fatmawati 2019; Naresh and Pandey 2009; Panja, Mondal, and Chattopadhyay 2016; Sota and Mogi 1989; Tapaswi, Ghosh, and Mukhopadhyay 1995; Zahid and Kribs 2021) |
| SEI | (n=3) | (Diallo et al. 2018; Ladreyt, Chevalier, and Durand 2022; Ladreyt et al. 2023) |
| ASI | (n=2) | (Ghassabzade and Bagherpoorfard 2021; Ndaïrou, Area, and Torres 2020) |
| I | (n=2) | (Kalita and Devi 2020a, 2020b) |

#### 3.4.3 Animal-Reservoir population

Most often, only one animal-reservoir population was described in models; however, the number of populations ranged up to six. Animals were also sometimes grouped as a “reservoir” representing a “pool of infection.” The most common group listed in models was pigs (n=19) followed by “reservoir” (n=7), cattle (n=2), chickens (n=2), dogs (n=2), ducks (n=2), “birds” (n=1) and “sows” (n=1).

Compartments used in models to reflect different animal-reservoir infection categories and the transition of the animal-reservoir populations over time were Maternal (M), Vaccinated (V), Susceptible (S), Exposed (E), Convalescent (C) and Recovered (R) (Table S2 for description of compartments). Models described the natural history of JEV in animal-reservoir populations using 13 different model structures and the most common was I (n=5) followed by SIRS (n=4) (Table 3).

**Table 3:** Number and types of model structures for animal-reservoir populations [Maternal (M), Vaccinated (V), Susceptible (S), Exposed (E), Convalescent (C) and Recovered (R)].

| Model structure | Animal types | Citation |
| --- | --- | --- |
| I (n=5) | Birds, pigs and reservoir | (Ghassabzade and Bagherpoorfard 2021; Goswami 2022; Kalita and Devi 2020a; Naresh and Pandey 2009; Ndaïrou, Area, and Torres 2020) |
| SIRS (n=4) | Reservoir | (Ghosh and Tapaswi 1999; Mukhopadhyay and Tapaswi 1994; Panja, Mondal, and Chattopadhyay 2016; Tapaswi, Ghosh, and Mukhopadhyay 1995) |
| SI (n=3) | Pigs | (Baniya and Keval 2021a; Dwivedi, Keval, and Baniya 2022; Kharismawati and Fatmawati 2019) |
| SIR (n=3) | Pigs | (Sota and Mogi 1989; Wada 1975; Zahid and Kribs 2021) |
| SIS (n=3) | Pigs | (Baniya and Keval 2021c, 2020b, 2021b) |
| MSEIR (n=1) | Cattle, chickens, dogs, ducks, pigs and sows | (Ladreyt, Chevalier, and Durand 2022) |
| MSIR (n=1) | Pigs | (Diallo et al. 2018) |
| MVSEIR (n=1) | Pigs | (Khan et al. 2014) |
| SEI (n=1) | Pigs | (Riad et al. 2017a) |
| SEICR (n=1) | Pigs | (Zhao et al. 2018) |
| SEIR (n=1) | Cattle, chickens, dogs, ducks and pigs | (Ladreyt et al. 2023) |
| VSI (n=1) | Pigs | (De et al. 2016) |
| VSIS (n=1) | Pigs | (Baniya and Keval 2020a) |

*3.5 State duration parameters and basic reproduction number*

### 3.5 State duration parameters and basic reproduction number

Descriptions of parameters and parameter values were clearly described in 21 models and the source of the data used to inform parameter values was clearly identified in 5 models. Units used for parameter values were not consistently identified. A total of 123 (human [n=23]; vector [n=45]; animal-reservoir [n=55]) unique parameters were identified over the 29 models. Of these parameters, 28% (n=34) were accompanied with sufficient information to extract units and values (human: 39% [n=9]; vector: 24% [n=11]; animal-reservoir: 25% [n=14]) (Figure 2; Tables S3, S4 and S5).

**Figure 2:**
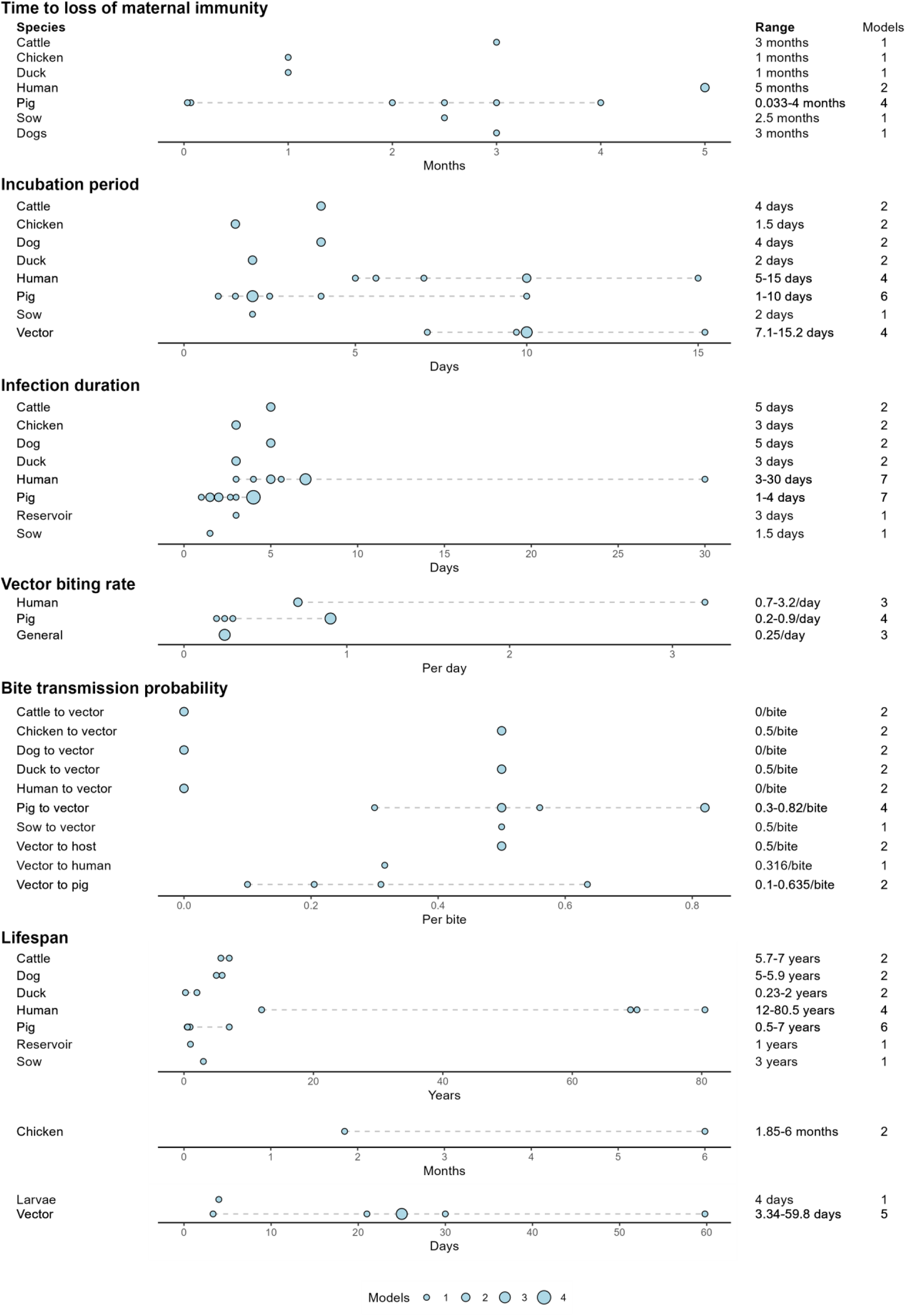
Parameter ranges (dotted lines) estimated from parameter values detailed in models when source of data was clearly identified. The size of the point indicates the number of models with a shared parameter value.

Basic reproduction number values were clearly described in 6 of the applied models. The basic reproduction number could be further broken down into three transmission types, vector-borne transmission, pig-to-pig transmission (vector free transmission), and combined vector-borne and pig-to-pig transmission (Figure 3; Table S6). The basic reproduction number range for pig-to-pig transmission was <1. When transmission type included vector-borne transmission, the basic reproduction number was >1 and up to 12, but more commonly between 1–3. One model investigated the impact of cattle on JEV transmission and estimated a basic reproduction number of 1.008 in the presence of cattle and 12.97 in the absence of cattle (Zahid and Kribs 2021).

**Figure 3:**
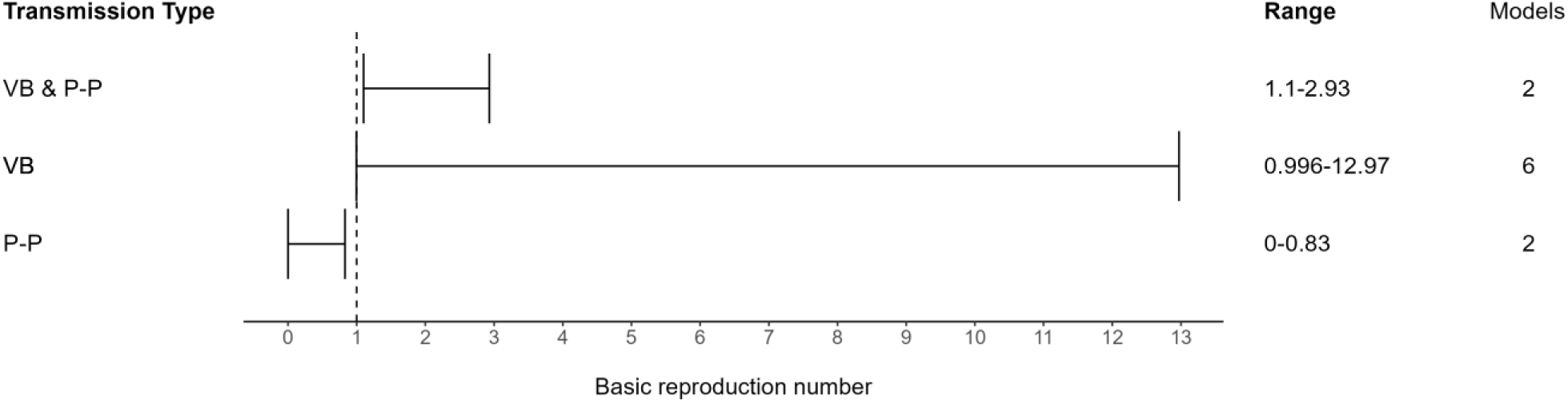
Basic reproduction number ranges estimated from 6 models using three transmission types. The total number of models using each transmission type is separately indicated. [*VB = vector-borne transmission; P-P = pig-to-pig transmission*]

### 3.6 Weather, location, and other factors

Eleven models included one or more parameters influenced by changes in weather or location, which in turn influenced their outputs. However, the information pertaining to weather and location often lacked specificity, with parameters either ambiguously defined or inadequately detailed. For instance, terms like “environmental discharges” were used in the models to describe environmental factors that contributed to the growth of reservoir and vector populations. Such discharges included a range of sources, such as household waste, open sewage draining, discarded tyres, and poorly ventilated houses.

The vector population size was influenced in all 11 models that considered environmental factors. Five models included a “vector carrying capacity”, which represents the maximum vector population size sustainable in a given environment (Baniya and Keval 2021c; Goswami 2022; Naresh and Pandey 2009; Ndaïrou, Area, and Torres 2020; Tapaswi, Ghosh, and Mukhopadhyay 1995). Weather variations influenced parameters in five models, leading to changes in vector population size (Ladreyt, Chevalier, and Durand 2022; Panja, Mondal, and Chattopadhyay 2016; Riad et al. 2017a; Riad et al. 2019; Zhao et al. 2018). Human-induced “environmental discharges” influenced parameters in three models, resulting in changes to the vector population size (Ghassabzade and Bagherpoorfard 2021; Naresh and Pandey 2009; Ndaïrou, Area, and Torres 2020) whilst the parameters of one model allowed for the vector population size to vary by geographic location (Riad et al. 2017a).

Three models were influenced by human-induced “environmental discharges”, similar to those impacting the vector population, which consequently led to changes in the animal-reservoir population size (Ghassabzade and Bagherpoorfard 2021; Naresh and Pandey 2009; Ndaïrou, Area, and Torres 2020). Furthermore, the parameters of one model varied based on the species and abundance of animals (birds), influenced by geographic location and time of year (Riad et al. 2017a). One model quantified animal (pig) population size by the daily consumption of pigs (Zhao et al. 2018). The authors also linked the decrease in pig abundance to a decrease in pig rearing licenses.

### 3.7 Prevention and control strategies

Twenty of the 29 models included parameters on prevention and control strategies with 18 models listing the influence of these parameters on model output. Seven models chose to apply prevention and control strategies through direct adjustment of the parameter that the strategy was anticipated to influence. Examples included reducing mosquito population size (insecticides), reducing bite and transmission rates (mosquito nets), reducing the size of the susceptible population (vaccinating the animal-reservoir or human populations), or reducing the growth of vector and animal-reservoir populations (by decreasing human induced “environmental discharges”) (Dwivedi, Keval, and Baniya 2022; Ghosh and Tapaswi 1999; Mukhopadhyay and Tapaswi 1994; Naresh and Pandey 2009; Riad et al. 2017a; Riad et al. 2019; Wada 1975).

In contrast, some authors explicitly modeled prevention and control strategies and included a specific parameter in the model to represent the strategy. Eleven models included parameters on the vaccination rate of susceptible humans or treatment rate of JEV infected humans (Baniya and Keval 2021c, 2020b, 2020a, 2021b; De et al. 2016; Dwivedi, Keval, and Baniya 2022; Goswami 2022; Kalita and Devi 2020b; Kharismawati and Fatmawati 2019; Mukhopadhyay and Tapaswi 1994; Panja, Mondal, and Chattopadhyay 2016), nine models included parameters on the vaccination rate of susceptible reservoir-animal populations or treatment of JEV infected reservoir-animal populations (Baniya and Keval 2020b, 2020a; De et al. 2016; Goswami 2022; Khan et al. 2014; Kharismawati and Fatmawati 2019; Panja, Mondal, and Chattopadhyay 2016; Wada 1975; Zhao et al. 2018), and four models included an insecticide control parameter which influenced the total mosquito population (De et al. 2016; Goswami 2022; Kharismawati and Fatmawati 2019; Panja, Mondal, and Chattopadhyay 2016). Six models evaluated the effectiveness and cost-effectiveness of various prevention and control strategies to recommend best approaches (Baniya and Keval 2020a; De et al. 2016; Goswami 2022; Kharismawati and Fatmawati 2019; Ladreyt, Chevalier, and Durand 2022; Panja, Mondal, and Chattopadhyay 2016). Lastly, two models assessed the impact of changing the proportions of competent and non-competent hosts within a community (Ladreyt, Chevalier, and Durand 2022; Zahid and Kribs 2021) and one explored the use of dogs as sentinel surveillance for JEV circulation (Ladreyt, Chevalier, and Durand 2022).

### 3.8 Identified limitations

Limitations were identified in 11 models. Authors of six records noted that their models did not include real-life variation that might influence outputs, such as the seasonality, heterogeneity and spatial distributions of populations, and inclusion of various JEV transmission scenarios (Diallo et al. 2018; Sota and Mogi 1989; Wada 1975; Zahid and Kribs 2021, 2021; Zhao et al. 2018). The authors of four records noted that the numbers in the field data used to validate their model might have been under-reported (for example, pig population data) or over-reported (for example, the use of JEV case data) (Khan et al. 2014; Ladreyt, Chevalier, and Durand 2022; Riad et al. 2017b, 2017a). Additionally, the authors of four models noted that there was limited information on contact structures between populations and there was limited host and vector attribute information, such as mosquito host feeding preferences, biting rates, and competency as a JEV vector. Authors also noted that parameters were made to fit the geographic scale of the model and that parameters were based on collected field data when an endemic state existed. Therefore, model parameters might not be appropriate when models are scaled up to cover a larger geographic region or when annual variations in JEV transmission occur (Diallo et al. 2018; Ladreyt, Chevalier, and Durand 2022; Sota and Mogi 1989). The epidemiology was also uncertain, such as the unknown impact that host species other than those commonly modeled can have in contributing to or limiting the spread of JEV or JEV introduction into susceptible populations (Diallo et al. 2018; Khan et al. 2014; Sota and Mogi 1989).

### 3.9 Sensitivity analyses

Sensitivity analysis was clearly described in nine models. The method used varied between models (normalized forward sensitivity index [n=6], next generation method [n=2] and Morris method [n=1]). Not all parameters were included in sensitivity analyses and they were selected based on the objectives and research question. The median number of parameters assessed was 8 (range: 1-51).

All nine models’ sensitivity analyses assessed the influence of input parameters on the basic reproduction number (Baniya and Keval 2020b, 2021b; Diallo et al. 2018; Dwivedi, Keval, and Baniya 2022; Goswami 2022; Kalita and Devi 2020a, 2020b; Ladreyt, Chevalier, and Durand 2022; Ladreyt et al. 2023). Most models assessed the impact of vector parameters on the basic reproduction number, finding that the vector biting rate, contact rate, death rate, population size, and probability of infection from a competent host were the most influential. Sensitivity analyses of two of these models quantified the influence of parameters and found that those related to the force of infection for vectors and hosts – specifically the vector biting rate, vector death rate, and the number of vectors – contributed the most to the total variance in basic reproduction number (Diallo et al. 2018; Ladreyt, Chevalier, and Durand 2022).

One model assessed the influence of input parameters on the maximum number of infectious pigs (Diallo et al. 2018). It found that the the number of vectors, the recovery rate of infected pigs, the rate of loss of maternal immunity of piglets, and their interactions, contributed the most to the total variance in the maximum number of infectious pigs.

## 4. Discussion

Models varied widely in their structure, including combinations of vector, animal-reservoir and human populations, with differing compartmental structures selected to describe aspects of the natural history of JEV infection in those populations. While this variation sometimes aligned with specific modeling goals, such as Zahid and Kribs (2021), in which the focus was on understanding the impact of cattle on joint occurrence of JE and leptospirosis, it also likely reflects great uncertainty about JEV epidemiology.

In their review on the ecology of JEV, Mulvey et al. (2021) noted the detection of JEV in various domestic animals and wildlife beyond ardeid wading birds and pigs. While pigs are recognised as amplifying hosts, overlooking other potential competent hosts may lead to underestimating the true extent of JEV transmission (Le Flohic et al. 2013; Levesque et al. 2024). Notably, JEV transmission in animal hosts has been observed to continue despite the phasing out of pig farming in Singapore over 30 years ago (Yap et al. 2019). Additionally, JEV has been associated with chicken density (Walsh et al. 2022). These observations suggest that other competent hosts, such as chickens, can sustain JEV circulation in regions with low pig densities, highlighting the potential importance of considering a broader range of hosts in understanding and controlling JEV transmission, especially in pig-free communities – this was reflected in few models in the current review, and non specifically investigated JEV circulation in species other than ardeid birds and pigs except Ladreyt et al. (2023) and Ladreyt, Chevalier, and Durand (2022).

Few of the modelling structures in the reviewed records allowed for simultaneous consideration of various factors influencing JEV spread, such as intrinsic incubation periods (for example in pigs) and extrinsic incubation periods (in vectors), while also enabling the assessment of different intervention strategies, like vaccination of pigs (Table 3). Despite the use of various compartmental structures and parameters, there was an absence of mosquito species-specific details in the vector populations in the models in the current review. Mosquito behaviour and ecological and host preferences vary between species known to transmit JEV (Zardini et al. 2024). Together with host competence, this variation explains the potential importance of the composition of animal populations in JEV transmission models.

Field data can inform model structure and parameter values, but the current review showed that such data were used in only 34% of the models. Integrating field data can enhance the accuracy of important parameter estimates which can improve model predictions (Grassly and Fraser 2008). Although this integration might overcome some epistemic uncertainty in model structure, epistemic uncertainty still existed with parameterisation because clarity regarding data sources and consistency in specifying units and time frames were limited. The identified discrepancies in parameter reporting led to challenges in model interpretation and comparison, highlighting the need for standardised reporting practices (Milwid et al. 2016; Garnett et al. 2011). Efforts to enhance transparency and consistency in parameterisation will be crucial for advancing the reliability and utility of JEV transmission models.

Six of the models based on field data described vector-borne basic reproduction numbers with wide variability which reflected differences in model structure and geographic origin of data, as well as factors like population density, seasonality, and vector mortality, that influence contact rates (Lord et al. 1996). Wide variability in basic reproduction numbers has also been observed in models of other diseases; for example, measles transmission, was represented with >20 different basic reproduction numbers, ranging from 5.4 to 18 (Guerra et al. 2017) and caution is warranted in interpreting these values beyond their region of calculation (Delamater et al. 2019). Although the reviewed models might not accurately reflect JEV transmission dynamics overall due to substantial differences in host and vector population structures across regions, calculating basic reproduction numbers in these models can help inform risk mitigation strategies. Changes in the basic reproduction number before and after interventions can indicate their effectiveness, even if the absolute predictions are not entirely accurate. For example, a significant reduction in basic reproduction number following an intervention suggests that the intervention is likely useful.

The authors of nine of the reviewed models conducted sensitivity analyses, mainly focusing on parameters influencing basic reproduction numbers. Notably, vector dynamics-related parameters like death rate, biting rate, and population size consistently emerged as significant influencers. Similar impacts have been observed in studies on diseases like African horse sickness (Lord et al. 1996) and malaria (Smith et al. 2007). Tennant and Recker (2018) highlighted the importance of obtaining field-relevant and species-dependent vector mortality rates for accurate modeling. However, most authors of reviewed models omitted sensitivity analysis, limiting readers’ understanding of parameter influences and potentially compromising the utility and robustness of model outputs.

The findings from models implementing prevention and control strategies underscore the complexities inherent in planning interventions for JEV transmission. While strategies targeting human populations successfully reduced human JE cases, those aimed at animal-reservoir and vector populations, such as pig vaccination and mosquito control, interrupted the JEV transmission cycle more comprehensively. The interconnectivity of model populations underscores the necessity for integrated prevention and control approaches. However, a gap remains in employing finer parameters to evaluate specific JEV control strategies and their effectiveness. This approach has been successfully used in dengue transmission models to assess their own control strategies (Ogunlade et al. 2023). For instance, instead of solely introducing a parameter to reduce mosquito abundance and inferring potential strategies like using insecticide sprays, models could offer a more nuanced understanding to enhance the precision of intervention strategies. Dengue transmission models have investigated the impact of chemical, biological, and environmental control methods to reduce mosquito numbers, as well as the long-term advantages and disadvantages of each. However, it is noteworthy that while dengue transmission is driven by only a few mosquito species (such as *Aedes aegypti* and *Aedes albopictus*) with highly specific habitat associations (Lambrechts, Scott, and Gubler 2010) JEV may have a greater number of mosquito species as potential vectors associated with a wider range of habitats (Van den Eynde et al. 2022).

The results of the 11 models that included the influence of environmental, weather, and geographic factors highlight the complex and multifaceted nature of JEV transmission. According to the model outputs, vector carrying capacity, weather variations, human-induced environmental discharges, and geographic location significantly impact vector and animal-reservoir abundance. Geographic and temporal variations have also been incorporated in other models to understand variations in mosquito-borne diseases (Caldwell et al. 2021), and human JE cases have been linked to changes in meteorological factors such as daily rainfall (Liu et al. 2020). Models incorporating environmental drivers of JEV transmission, particularly climate features, might more accurately predict JE occurrence.

The limitations highlighted by the authors of the reviewed models re-iterated the challenges in JEV transmission modeling that are identified above. Several models overlooked factors such as seasonal and spatial impacts, population heterogeneity, and diverse JEV transmission scenarios. Authors also expressed concerns regarding the accuracy of field data used for model validation, citing instances of both under and overreporting of data, highlighting the need for reliable and more accurate data sources. Addressing these limitations is essential for refining model inputs and advancing future JEV transmission models.

## 5. Conclusions

Overall, this review provides insight into the literature on JEV transmission modelling, revealing both progress and challenges in understanding and mitigating the impact of JE occurrence. Despite the significant global burden of JE, it is notable that only a limited number of models have been developed to study the viral transmission dynamics, indicating a gap in our understanding of current prevention and control strategies, as well as preparedness for JEV emergence in new regions. Increased investment in JEV transmission modelling is essential to develop robust tools that can inform decision-making in JEV prevention and control to work towards reducing the global burden of JEV and safeguarding the health of populations at risk.

## Data availability

All data supporting the findings of this review are included in the main article and the supplementary materials.

## Authors’ contributions

*Conceptualisation*: Victoria J. Brookes, Troy A. Laidlow; *Data curation*: Troy A. Laidlow; *Format analysis*: Troy A. Laidlow; *Funding acquisition*: Victoria J. Brookes, Troy A. Laidlow; *Investigation*: All authors; *Project administration*: Troy A. Laidlow; *Resources*: Troy A. Laidlow; *Supervision*: Victoria J. Brookes, Kerrie E. Wiley; Ruth N. Zadoks; *Validation*: All authors; *Visualisation*: Troy A. Laidlow; *Writing – original draft*: Troy A. Laidlow; *Writing – review & editing*: All authors.

## Acknowledgements

We thank the librarians at The University of Sydney for their assistance with this study.

## Supplementary materials

*Supplementary 1* includes Figure S1: Distribution of records published in conference proceedings and journals; Figure S2: Distribution of records published by year of publication Figure S3: Countries where Japanese encephalitis virus has been identified and countries from which field data were obtained to be used in Japanese encephalitis virus disease transmission models; Table S1: Forms used at each level of the scoping review; Table S2: Disease compartments and use within model structures; Table S3, S4, S5, S6: Model parameters and value ranges; and Table S6: Identified basic reproduction numbers.

