## Supplementary Material 1 for "Scoping review of Japanese encephalitis virus transmission models"

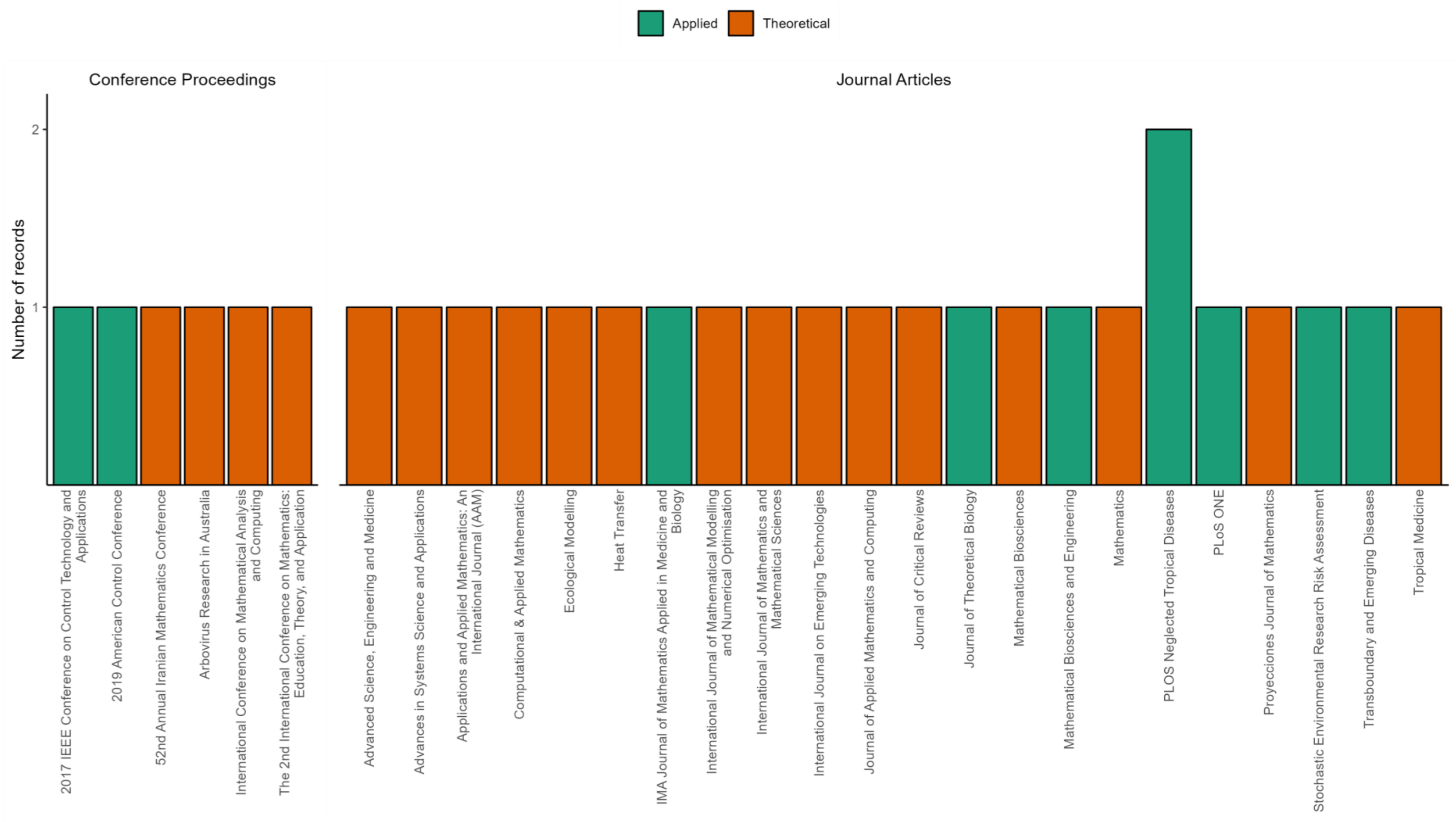


Figure S1: Distribution of peer-reviewed records published from 1975–2023 (inclusive) in conference proceedings (n=6) and journals (n=22), categorised by source, stratified by application of model to real Japanese encephalitis virus transmission settings (n=10 applied, n=19 theoretical).

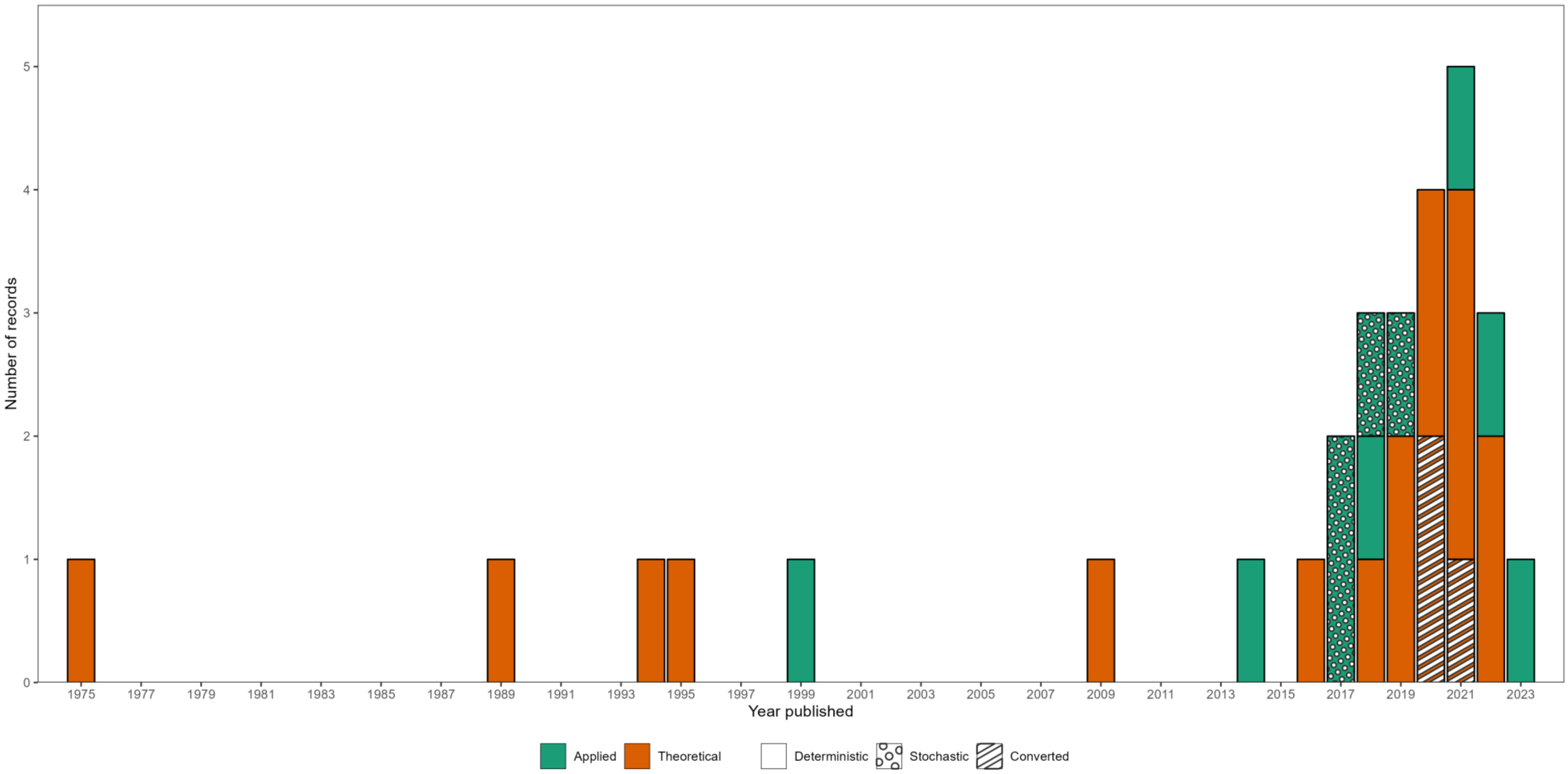


Figure S2: Distribution of records published from 1975–2023 (inclusive) by year of publication, application of model to real Japanese encephalitis virus transmission setting (n=10 applied, n=19 theoretical), and model classification as deterministic (n=22), stochastic (n=4) or converted (n=3).

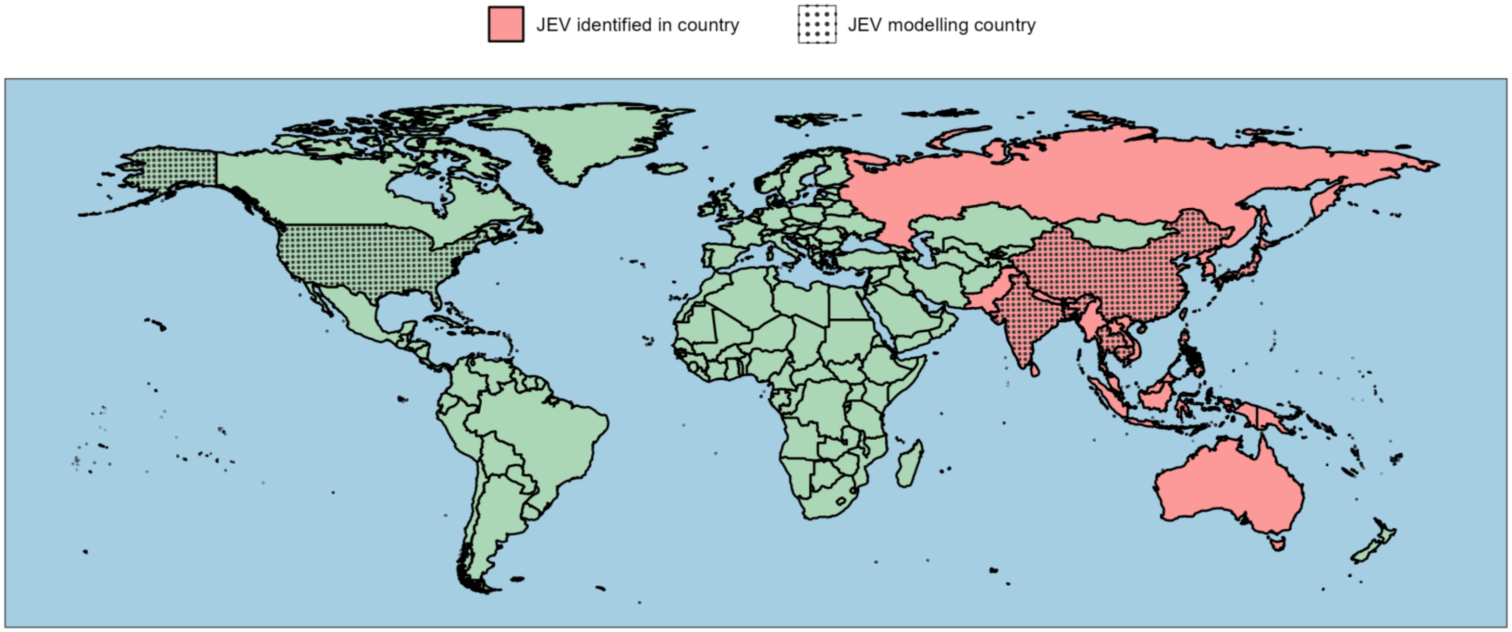


| Global region | | Country & external territories (subregions if applicable) |
|---|---|---|
| Western Pacific | (n=6) | Cambodia (Phnom Penh & Kandal province [Khsach Kandal, Kien Svay, and Kaoh Thum]), China (Hong Kong), Japan (Saitama prefecture), Philippines, and Taiwan |
| South-East Asia | (n=3) | Bangladesh (Rajshahi, Nawabgonj, and Naogaon), India (Odisha), and Thailand |
| African | (n=1) | French Overseas Département of Réunion |
| Americas | (n=1) | United States of America (Florida, North Carolina, and South Carolina) |

Figure S3: Countries where Japanese encephalitis virus has been identified (Centers for Disease Control and Prevention 2023) and countries from which field data were obtained to be used in Japanese encephalitis virus disease transmission models. The table below complements the map by detailing World Health Organization regions, the number of models per region, and country and external territories (including subregions where applicable) where data informed the models, noting instances where data collection and model development occurred in the same country.

Table S1: Forms used at each level of the scoping review

*Level 1: Screening on title and abstract*

| **Question** | **Inclusion** |
|---|---|
| Could this record be about the design, development, and/or implementation of a population-based JEV transmission model? | Yes |
| Are the title and abstract in English? | Yes |
| Could this record be peer-reviewed PRIMARY literature? | Yes |
| Include this article? | Yes |

*Level 2: Screening on full record*

| **Question** | **Inclusion** |
|---|---|
| Is this record about the design, development, and/or implementation of a population-based JEV transmission model | Yes |
| Is all of the content of this record in English? | Yes |
| Is this record peer-reviewed PRIMARY literature? | Yes |
| Include this article? | Yes |

*Level 3: Data charting*

| **Identification** |
|---|
| Article reference |
| **Background** |
| Is this article part of conference proceedings? |
| What is the name of the journal the article is in? / What is the name of the conference proceedings? |
| What year was the study published? |
| Is the article theoretical or applied? |
| What are the aims/objectives of the model? |

| Applied Models |
| --- |
| If the model is applied, what is the year range used in the model? |
| If the model is applied, what is the duration of model simulations? |
| If the model is applied, what is the geographic location used in model — global region? |
| --- Choice: African, Eastern Mediterranean, European, Americas, South-East Asia, and Western Pacific |
| If the model is applied, what is the geographic location used in model — country? |
| If the model is applied, what is the geographic location used in model — country subregion? |
| If the model is applied, what is the geographic location used in the model specific to country subregion, country, global region, or combination? |
| **Model Structure** |
| What type of model is being used? |
| --- Choice: Deterministic, Stochastic |
| What time step is being used in the model? |
| --- Choice: Difference, Differential, Discrete time, Continuous time |
| Have animals (non-human mammals and birds) been modelled? |
| What are the details of the animals (non-human mammals and birds) being modelled? |
| Have vectors been modelled? |
| What are the details of the vectors being modelled? |
| How many populations are represented in the model? |
| What are the compartments of the human population in the model? |
| What is the structure of the human population in the model? |
| What are the parameters used in the human population? |
| What are the compartments of the vector population in the model? |
| What is the structure of the vector population in the model? |
| What are the parameters used for the vector population? |
| What are the compartments of the animal (non-human mammals and birds) population in the model? |
| What is the structure of the animal (non-human mammals and birds) population in the model? |
| What are the parameters used for the animal (non-human mammals and birds) population? |
| What is the contact mix of the model? |
| What are the details of the contact mix in the model? |

| |
|---|
| **Applied Models** |
| Have the model populations been stratified into groups? |
| What are the details of the stratified groups? |
| **Data Source** |
| What parameters used in the model are hypothetical? |
| What parameters used in the model are from literature? |
| What parameters used in the model are from data collected by authors (empirical data)? |
| What parameters used in the model are from fitting the model? |
| **Other Variables** |
| Have weather conditions been modelled? |
| What are the details of the weather conditions that have been modelled? |
| Was there ecological data (other than weather) used in the model? |
| What are the details of the ecological data (other than weather) that have been modelled? |
| Were there socio-economic factors used in the model? |
| What are the details of the socio-economic data that have been modelled? |
| Are there prevention/control measures in the model? |
| What are the details of the prevention/control measures that have been modelled? |
| What are the model outputs? |
| Was sensitivity analysis performed on the model? |
| What are the details and outputs of the sensitivity analysis? |
| **Limitations** |
| Are there any limitations identified by the author? |
| What are the details and limitations identified by the author? |

Table S2: Compartment, compartment abbreviations, and use within model structures for description of Japanese encephalitis virus (JEV) dynamics in populations.

| Compartment | Abbreviation | Description |
|---|---|---|
| Aquatic | A | *Vector population*: Individuals in the early growing phase (egg, larvae, and pupae stages) who are not at risk of being infected with JEV. |
| Maternal | M | Individuals who have passive immunity due to maternal antibodies. |
| Vaccinated | V | Individuals who have immunity due to vaccination. |
| Susceptible | S | Individuals who are not infected but are at risk of becoming infected with JEV. |
| Exposed | E | Individuals who have been infected with JEV but are not yet infectious. |
| Infected | I | Individuals who are infected and infectious (capable of transmitting JEV to other individuals) |
| Convalescent | C | *Pig population*: Individuals who are infected and infectious (capable of transmitting JEV to other individuals) via oronasal secretions only. |
| Recovered | R | Individuals who have recovered from JEV infection and have gained immunity and are no longer susceptible to infection. |

Table S3: Model parameters and value ranges (when records provided sufficient information) used in representing Japanese encephalitis virus disease transmission in human populations.

| **Parameter** | **Value Range** | **Citation** |
|---|---|---|
| bite transmission probability (vector-human) | 0.316 per | (Zahid and Kribs 2021) |
| time to death (infected) | 26-120 days | (Ghosh and Tapaswi 1999; Zahid and Kribs 2021) |
| lifespan | 12-80.5 years | (Ghosh and Tapaswi 1999; Ladreyt, Chevalier, and Durand 2022; Ladreyt et al. 2023; Zahid and Kribs 2021) |
| effective contact rate (reservoir-human) | 0.001-21 | (Ghosh and Tapaswi 1999; Riad et al. 2019) |
| incubation period | 5-15 days | (Ladreyt, Chevalier, and Durand 2022; Ladreyt et al. 2023; Riad et al. 2017b, 2019) |
| infection duration | 3-30 days | (Baniya and Keval 2020b; Ghosh and Tapaswi 1999; Ladreyt, Chevalier, and Durand 2022; Ladreyt et al. 2023; Riad et al. 2017b, 2019; Zahid and Kribs 2021) |
| spill-over rate | 5-13 days | (Zhao et al. 2018) |
| time to loss of maternal immunity | 5 months | (Ladreyt, Chevalier, and Durand 2022; Ladreyt et al. 2023) |

Table S4: Model parameters and value ranges (when records provided sufficient information) used in representing Japanese encephalitis virus disease transmission in vector populations.

| **Parameter** | **Value Range** | **Citation** |
|---|---|---|
| bite transmission probability (cattle-vector) | 0 per bite | (Ladreyt, Chevalier, and Durand 2022; Ladreyt et al. 2023) |
| bite transmission probability (chicken-vector) | 0.5 per bite | (Ladreyt, Chevalier, and Durand 2022; Ladreyt et al. 2023) |
| bite transmission probability (dog-vector) | 0 per bite | (Ladreyt, Chevalier, and Durand 2022; Ladreyt et al. 2023) |
| bite transmission probability (duck-vector) | 0.5 per bite | (Ladreyt, Chevalier, and Durand 2022; Ladreyt et al. 2023) |
| bite transmission probability (human-vector) | 0 per bite | (Ladreyt, Chevalier, and Durand 2022; Ladreyt et al. 2023) |
| bite transmission probability (pig-vector) | 0.3-0.82 per bite | (Diallo et al. 2018; Ladreyt, Chevalier, and Durand 2022; Ladreyt et al. 2023; Zahid and Kribs 2021) |
| bite transmission probability (sow-vector) | 0.5 per bite | (Ladreyt, Chevalier, and Durand 2022) |
| bite transmission probability (vector-host) | 0.5 per bite | (Ladreyt, Chevalier, and Durand 2022; Ladreyt et al. 2023) |
| biting rate | 0.25 per day | (Ladreyt, Chevalier, and Durand 2022; Ladreyt et al. 2023; Zahid and Kribs 2021) |
| biting rate (human) | 0.7-3.2 per day | (Baniya and Keval 2020b; De et al. 2016; Kharismawati and Fatmawati 2019) |
| biting rate (pig) | 0.2-0.9 per day | (Baniya and Keval 2020b; De et al. 2016; Diallo et al. 2018; Kharismawati and Fatmawati 2019) |
| contact rate (vector-human) | 771250 bites/day | (Zahid and Kribs 2021) |
| contact rate (vector-pig) | 142102 bites/day | (Zahid and Kribs 2021) |
| lifespan | 3.34-59.8 days | (Diallo et al. 2018; Ladreyt, Chevalier, and Durand 2022; Ladreyt et al. 2023; Ndaïrou, Area, and Torres 2020; Zahid and Kribs 2021) |
| lifespan (larvae) | 4 days | (Ndaïrou, Area, and Torres 2020) |
| extrinsic incubation period | 7.1-15.2 days | (Diallo et al. 2018; Ladreyt, Chevalier, and Durand 2022; Ladreyt et al. 2023; Wada 1975) |
| feeding preference (pig-cattle) | 1:1.7 pig:cattle | (Ladreyt, Chevalier, and Durand 2022; Ladreyt et al. 2023) |

| Parameter | Value Range | Citation |
|---|---|---|
| feeding preference (pig-chicken) | 1:0.09 pig:chicken | (Ladreyt, Chevalier, and Durand 2022; Ladreyt et al. 2023) |
| feeding preference (pig-dog) | 1:0.12 pig:dog | (Ladreyt, Chevalier, and Durand 2022; Ladreyt et al. 2023) |
| feeding preference (pig-duck) | 1:0.43 pig:duck | (Ladreyt, Chevalier, and Durand 2022; Ladreyt et al. 2023) |
| feeding preference (pig-human) | 1:0.5 pig:human | (Ladreyt, Chevalier, and Durand 2022; Ladreyt et al. 2023) |
| feeding preference (pig-pig) | 1:1 pig:pig | (Ladreyt, Chevalier, and Durand 2022; Ladreyt et al. 2023) |
| feeding preference (pig-sow) | 1:1 pig:sow | (Ladreyt, Chevalier, and Durand 2022) |
| recruitment duration (eggs) | 1.67 days | (Ndaïrou, Area, and Torres 2020) |
| vertical transmission probability | 0.04 | (Zahid and Kribs 2021) |

Table S5: Model parameters and value ranges (when records provided sufficient information) used in representing Japanese encephalitis virus disease transmission in animal-reservoir populations.

| **Parameter** | **Value Range** | **Citation** |
|---|---|---|
| bite transmission probability (reservoir-vector) | 0.00021 per bite | (Ndaïrou, Area, and Torres 2020) |
| bite transmission probability (vector-pig) | 0.1-0.635 per bite | (Diallo et al. 2018; Zahid and Kribs 2021) |
| convalescent duration (pig) | 1-4 days | (Zhao et al. 2018) |
| lifespan (cattle) | 5.7-7 years | (Ladreyt, Chevalier, and Durand 2022; Ladreyt et al. 2023) |
| lifespan (chicken) | 1.85-6 months | (Ladreyt, Chevalier, and Durand 2022; Ladreyt et al. 2023) |
| lifespan (dog) | 5-5.9 years | (Ladreyt, Chevalier, and Durand 2022; Ladreyt et al. 2023) |
| lifespan (duck) | 0.23-2 years | (Ladreyt, Chevalier, and Durand 2022; Ladreyt et al. 2023) |
| lifespan (pig) | 0.5-7 years | (Khan et al. 2014; Ladreyt, Chevalier, and Durand 2022; Ladreyt et al. 2023; Wada 1975; Zahid and Kribs 2021; Zhao et al. 2018) |
| lifespan (reservoir) | 1 years | (Ghosh and Tapaswi 1999) |
| lifespan (sow) | 3 years | (Ladreyt, Chevalier, and Durand 2022) |
| effective contact rate (vector-pig) | 0-0.4 | (Khan et al. 2014; Zhao et al. 2018) |
| effective contact rate (vector-reservoir) | 0.55-0.6 | (Ghosh and Tapaswi 1999) |
| external introduction proportion (imported pig) | 0.43-1.45 | (Zhao et al. 2018) |
| external introduction proportion (other hosts) | 0.05 | (Khan et al. 2014) |
| incubation period (cattle) | 4 days | (Ladreyt, Chevalier, and Durand 2022; Ladreyt et al. 2023) |
| incubation period (chicken) | 1.5 days | (Ladreyt, Chevalier, and Durand 2022; Ladreyt et al. 2023) |
| incubation period (dog) | 4 days | (Ladreyt, Chevalier, and Durand 2022; Ladreyt et al. 2023) |

| Parameter | Value Range | Citation |
| --- | --- | --- |
| incubation period (duck) | 2 days | (Ladreyt, Chevalier, and Durand 2022; Ladreyt et al. 2023) |
| incubation period (pig) | 1-10 days | (Khan et al. 2014; Ladreyt, Chevalier, and Durand 2022; Ladreyt et al. 2023; Riad et al. 2017a; Wada 1975; Zhao et al. 2018) |
| incubation period (sow) | 2 days | (Ladreyt, Chevalier, and Durand 2022) |
| infection duration (cattle) | 5 days | (Ladreyt, Chevalier, and Durand 2022; Ladreyt et al. 2023) |
| infection duration (chicken) | 3 days | (Ladreyt, Chevalier, and Durand 2022; Ladreyt et al. 2023) |
| infection duration (dog) | 5 days | (Ladreyt, Chevalier, and Durand 2022; Ladreyt et al. 2023) |
| infection duration (duck) | 3 days | (Ladreyt, Chevalier, and Durand 2022; Ladreyt et al. 2023) |
| infection duration (pig) | 1-4 days | (Diallo et al. 2018; Khan et al. 2014; Ladreyt, Chevalier, and Durand 2022; Ladreyt et al. 2023; Wada 1975; Zahid and Kribs 2021; Zhao et al. 2018) |
| infection duration (reservoir) | 3 days | (Ghosh and Tapaswi 1999) |
| infection duration (sow) | 1.5 days | (Ladreyt, Chevalier, and Durand 2022) |
| time to loss of maternal immunity (cattle) | 3 months | (Ladreyt, Chevalier, and Durand 2022) |
| time to loss of maternal immunity (chicken) | 1 months | (Ladreyt, Chevalier, and Durand 2022) |
| time to loss of maternal immunity (dogs) | 3 months | (Ladreyt, Chevalier, and Durand 2022) |
| time to loss of maternal immunity (duck) | 1 months | (Ladreyt, Chevalier, and Durand 2022) |
| time to loss of maternal immunity (pig) | 0.033-4 months | (Diallo et al. 2018; Khan et al. 2014; Ladreyt, Chevalier, and Durand 2022; Wada 1975) |
| time to loss of maternal immunity (sow) | 2.5 months | (Ladreyt, Chevalier, and Durand 2022) |

Table S6: Identified basic reproduction numbers and value ranges (when records provided sufficient information) used in representing Japanese encephalitis virus disease transmission.

| Types of transmission | Value Range | Citation |
|---|---|---|
| Pig-pig | 0-0.83 | (Diallo et al. 2018; Zhao et al. 2018) |
| Vector-borne & pig-pig | 1.1-2.93 | (Diallo et al. 2018; Zhao et al. 2018) |
| Vector-borne | 0.996-12.97 | (Diallo et al. 2018; Khan et al. 2014; Ladreyt, Chevalier, and Durand 2022; Riad et al. 2017b; Zahid and Kribs 2021; Zhao et al. 2018) |
